# Tissue-Specific Regulation of Postprandial Non-Esterified Fatty Acid Metabolism in individuals with prediabetes

**DOI:** 10.64898/2026.10.23.26365753

**Authors:** Elisa Le Roux, Sophie Hogan-Lamarre, Marie Lepitre, Frédérique Frisch, Lauralyne Dumont, Mélanie Fortin, Gabriel Richard, Christophe Noll, Otman Sarrhini, Warner Mampuya, Serge Phoenix, Lucie Bouffard, Brigitte Guérin, Éric E. Turcotte, Femke Rutters, Parminder Raina, Jean-Pierre Desprès, Joris Hoeks, Patrick Schrauwen, André C. Carpentier, Denis P. Blondin

## Abstract

After a meal, it is generally assumed that tissue non-esterified fatty acids (NEFA) uptake falls in proportion of the reduction of systemic NEFA appearance from insulin-mediated suppression of adipose tissue lipolysis. Here, we combined dynamic [^11^C]palmitate PET/CT, stable isotope tracers, and MRI to measure palmitate uptake, oxidation, and esterification in the heart, liver, skeletal muscle, and subcutaneous and visceral adipose tissue, in the fasted state and 60min after a mixed meal in individuals with prediabetes. Net palmitate uptake decreased in all tissues, but most tissues extracted a larger fraction of circulating palmitate after the meal. The intracellular fate of palmitate differed between organs. The heart shifted toward oxidation, the liver toward esterification, maintaining VLDL-palmitate release, and this shift was smaller in individuals with more liver fat. Both adipose depots increased fractional esterification, and visceral fat almost stopped releasing palmitate. Thus, plasma NEFA overestimates the postprandial reduction in lean tissue NEFA exposure.

**Highlights:**

- After a meal, net palmitate uptake fall in all tissues but extraction rises in most
- After a meal, the liver directs palmitate toward esterification and the heart toward oxidation
- Plasma NEFA level underestimate lean tissue NEFA exposure after a meal in prediabetes

## 1 Introduction

Adipose tissue plays a critical role in regulating lipid metabolism and whole-body energy homeostasis. In the fasted state, adipose tissue lipolysis from adipose triglyceride lipase (ATGL) and hormone-sensitive lipase (HSL) maintains elevated circulating concentrations of non-esterified fatty acids (NEFA)^1^, which constitute an important energy substrate for lean tissues. Following the ingestion of a mixed meal, insulin suppresses adipose tissue lipolysis through inhibition of ATGL-and HSL-mediated triglyceride hydrolysis^2^, thereby markedly decreasing the systemic appearance of NEFA. Insulin further enhances fatty acid esterification in adipocytes by increasing glucose uptake to generate the glycerol precursor, glycerol-3-phosphate, required for triglyceride formation and stimulating diacylglycerol acyl-transferase activity^3–5^. Together, these insulin-driven changes reduce NEFA release while promoting their re-esterification and storage in adipose tissue during the postprandial period.

Although the postprandial fall in circulating NEFA concentrations is well established, less is known about the proportional handling of NEFA across different tissues and their intracellular fate following a meal. Organ-specific NEFA uptake is a function of both plasma concentration and the fractional extraction. If fractional extraction is assumed constant after a meal, tissue NEFA uptake would be expected to fall in proportion to plasma NEFA concentrations. At the whole-body level, however, plasma NEFA clearance is inversely related to the systemic rate of appearance of NEFA in healthy men^6^. Studies using arterial-venous concentration differences and stable isotope labelling have provided important insights into the regulation of NEFA uptake under insulin stimulation in the postprandial state in skeletal muscle and adipose tissue^7–9^. Whole-body measurements, arterial-venous approaches, and stable isotope tracer methodologies have provided important additional insights into systemic and tissue-level NEFA fluxes, including tissue uptake and storage^10–12^, but they have largely focused on subcutaneous adipose tissue alone^6,8,13,14^. Furthermore, these methods cannot determine lean tissue NEFA oxidation and esterification rates. Thus, postprandial changes in lean tissue NEFA uptake and intracellular metabolic partitioning remain unclear.

The postprandial handling of NEFA is particularly relevant in prediabetes since lipid metabolism is already altered before overt type 2 diabetes develops. Indeed, in prediabetes, adipose tissue presents reduced catecholamine-stimulated lipolysis, blunted insulin suppression of lipolysis and increased trapping of dietary fatty acids, affecting both lipid release and storage^14–16^. When adipose tissue fails to buffer the postprandial flux of fatty acids, more NEFA are delivered to lean tissues, where they can accumulate as ectopic lipid and impair insulin action^10,17^. Whether the fasting-to-postprandial transition induces coordinated or tissue-specific changes in NEFA uptake and downstream metabolism across lean tissues and adipose depots therefore remains unknown. Identifying which tissues take up NEFA after a meal, their intracellular metabolic fate, could help explain how ectopic lipid accumulates as prediabetes progresses and identify tissue-specific targets for the prevention of type 2 diabetes.

We therefore aimed to compare fasting and early-postprandial NEFA metabolism in skeletal muscle, liver, heart, subcutaneous adipose tissue (SAT), and visceral adipose tissue (VAT) in individuals with prediabetes, by combining [^11^C]palmitate positron emission tomography with computed tomography (PET/CT), magnetic resonance imaging (MRI), and stable isotope tracers. We also examined whether the fasting-to-postprandial changes in tissue-specific palmitate metabolism were associated with changes in circulating insulin levels, glucose and NEFA kinetics, anthropometric markers of adiposity, and organ-specific lipid content. We hypothesized that the early postprandial state, at the peak insulin and at the nadir of NEFA concentrations, would shift NEFA partitioning toward uptake and esterification in adipose tissue. Because adiposity and ectopic lipid deposition are associated with insulin resistance, we further hypothesized that organ-specific palmitate flux would be associated with the magnitude of the insulin response, NEFA and glucose kinetics, adiposity and ectopic fat content.

## 2 Material and methods

### 2.1 Participants and Ethical Approval

Participants were recruited from the local community through posters, social media posts, newspaper advertisements, and clinical records available through the *Données Organisées du Réseau Informatique de Santé de l’Estrie* (DORISE). Overall, 56 individuals were screened for eligibility. Among these, 15 individuals were excluded because they did not meet the inclusion criteria, and 6 were excluded due to claustrophobia, back pain, or due to availability. A total of 35 participants with prediabetes (24 women and 11 men) were enrolled in the study. All participants provided written consent after being informed about the experimental procedures. The study protocol adhered to the Declaration of Helsinki and was approved by the Human Ethics Committee of the Centre de recherche du Centre Hospitalier Universitaire de Sherbrooke (MP-31-2021-401). The data presented in this manuscript are a secondary analysis of a broader trial registered at ClinicalTrials.gov (NCT05123963).

Participants aged 45 to 75 years with a BMI >25 kg·m^-2^ and prediabetes were included. Prediabetes was defined following Diabetes Canada criteria, by at least one of the following criteria: (1) Fasting glucose between 6.1 and 6.9 mmol·L^-1^; (2) 2-h glycemia post-75 g oral glucose tolerance test (OGTT) between 7.8 mmol·L^-1^ and 11.0 mmol·L^-1^, with anHbA1c between 6.0 and 6.4%; (3) Glucose clearance rate ≤360 mL·kg^-1^·min^-1^ as determined using the oral glucose insulin sensitivity index (OGIS)^18^. Participant characteristics are provided in **Table 1**.

**Table 1:** Participants’ characteristics.

|  | Women (n=24) | Men (n=11) | All (n=35) |
| --- | --- | --- | --- |
|  | Mean [95%CI] | Mean [95%CI] | Mean [95%CI] |
| *Anthropometry and body composition* |  |  |  |
| **Age (years)** | 58 [55-61] | 62 [57-67] | 59 [57-62] |
| **Body mass (kg)** | 83.0 [76.4-89.5] | 97.5 [86.2-108.9] | 87.6 [81.7-93.4] |
| **BMI (kg·m<sup>-2</sup>)** | 31.5 [29.1-33.9] | 32.5 [28.9-36.1] | 31.8 [29.9-33.7] |
| **Lean mass (kg)** | 40.1 [37.7-42.5] | 57.4 [53.0-61.8] | 45.5 [42.1-49.0] |
| **LMI (kg·m<sup>-2</sup>)** | 15.2 [14.4-16.0] | 19.1 [17.7-20.5] | 16.4 [15.5-17.3] |
| **Fat mass (kg)** | 41.3 [35.9-46.7] | 36.1 [28.6-43.6] | 39.7 [35.4-43.9] |
| **FMI (kg·m<sup>-2</sup>)** | 15.7 [13.6-17.8] | 12.0 [9.5-14.4] | 14.5 [12.9-16.2] |
| **Waist circumference (cm)** | 97 [93-102] | 108 [101-114] | 101 [97-105] |
| *Glycemic control<sup>b</sup>* |  |  |  |
| **HbA1c (%)** | 5.7 [5.5-5.8] | 5.7 [5.4-5.9] | 5.7 [5.5-5.8] |
| **Fasting glucose (mmol·L<sup>-1</sup>)** | 6.1 [5.8-6.3] | 6.2 [5.9-6.4] | 6.1 [5.9-6.3] |
| **2-h post-OGTT glucose (mmol·L<sup>-1</sup>)** | 8.5 [7.2-9.7] | 8.4 [7.2-9.6] | 8.4 [7.5-9.3] |
| **OGIS (mL·kg<sup>-1</sup>·min<sup>-1</sup>)** | 297 [275-318] | 289 [268-310] | 294 [278-310] |
| *Tissues PDFF or attenuation* |  |  |  |
| **Myocardium (HU)** | 30 [27-33] | 32 [27-37] | 30 [28-33] |
| **Skeletal muscle (%)** | 13.6 [12.2-15.0] | 10.7 [8.5-13.0] | 12.7 [11.5-14.0] |
| **Liver<sup>a</sup> (%)** | 8.6 [5.9-22.7] | 10.9 [7.6-18.8] | 9.0 [6.0-20.2] |
| *Adipose tissue lipid unsaturation* |  |  |  |
| **Subcutaneous AT NDB** | 2.62 [2.55-2.70] | 2.62 [2.58-2.65] | 2.62 [2.57-2.67] |
| **Visceral AT NDB** | 2.60 [2.53-2.67] | 2.70 [2.58-2.83] | 2.63 [2.57-2.69] |
| *Index of tissue insulin resistance* |  |  |  |
| **Adipose tissue insulin resistance<sup>a</sup> (AU)** | 115 [34-155] | 66 [50-144] | 75 [45-156] |
| **Liver insulin resistance<sup>a</sup> (AU)** | 256 [181-595] | 228 [188-258] | 251 [181-418] |

<sup>a</sup>Data are median [Q1-Q3] because of their skewed distribution. <sup>b</sup>Data are from inclusion visit. AT, adipose tissue; BMI, body mass index; FMI, fat mass index; LMI, lean mass index; NDB, number of double bonds; OGIS, oral glucose insulin sensitivity index. Myocardium attenuation is expressed in Hounsfield unit; adipose tissue and liver insulin resistance are expressed in arbitrary unit.

Exclusion criteria were cardiovascular disease based on medical history, physical examination, or abnormal electrocardiogram (history of ischemic heart disease, tachyarrhythmia, QT interval prolongation or use of medication known to prolong the QT interval; pacemaker); treatment with medication known to affect lipid or carbohydrate metabolism, including GLP-1 receptor agonists and menopausal hormone therapy, except statins (stopped 3 weeks before metabolic visit), metformin, and antihypertensive medication (stopped 7 days before metabolic visit); liver or kidney disease other than uncomplicated metabolic dysfunction-associated steatotic liver disease (MASLD) or isolated mild proteinuria; uncontrolled thyroid disorder, previous pancreatitis, bleeding disorder, or other major illness; severe uncontrolled hypertension (systolic blood pressure ≥180 mmHg or diastolic pressure ≥110 mmHg); prior history of, or current fasting plasma cholesterol level >7 mmol.L^-1^ or triglycerides >6 mmol.L^-1^; severe gastrointestinal disorders; smoking (>1 cigarette/day) or alcohol consumption >2 drinks/day; PET or CT scan within the previous year; contraindication to discontinuation of statins for 3 weeks or metformin or antihypertensive medication for 7 days; blood donation two months before the study; or inability to give informed consent.

### 2.2 Overall Study Design

The study was conducted at the Centre de Recherche du Centre Hospitalier Universitaire (CRCHUS of Sherbrooke, Québec, Canada. Before the metabolic study visit, participants followed a 3-day isocaloric diet (33% fat, 17% protein, 50% carbohydrates) based on estimated energy expenditure^19^. After an overnight fast, body mass was measured with a calibrated scale (TBF 300 A, Tanita Corporation of America Inc., Arlington Heights, IL), and waist circumference (WC) was measured. Fat mass (FM) and lean mass (LM) were assessed by dual energy X-ray absorptiometry (DXA, HOLOGIC QDR 4500W). Study design is depicted in **Figure 1**.

**Figure 1:**
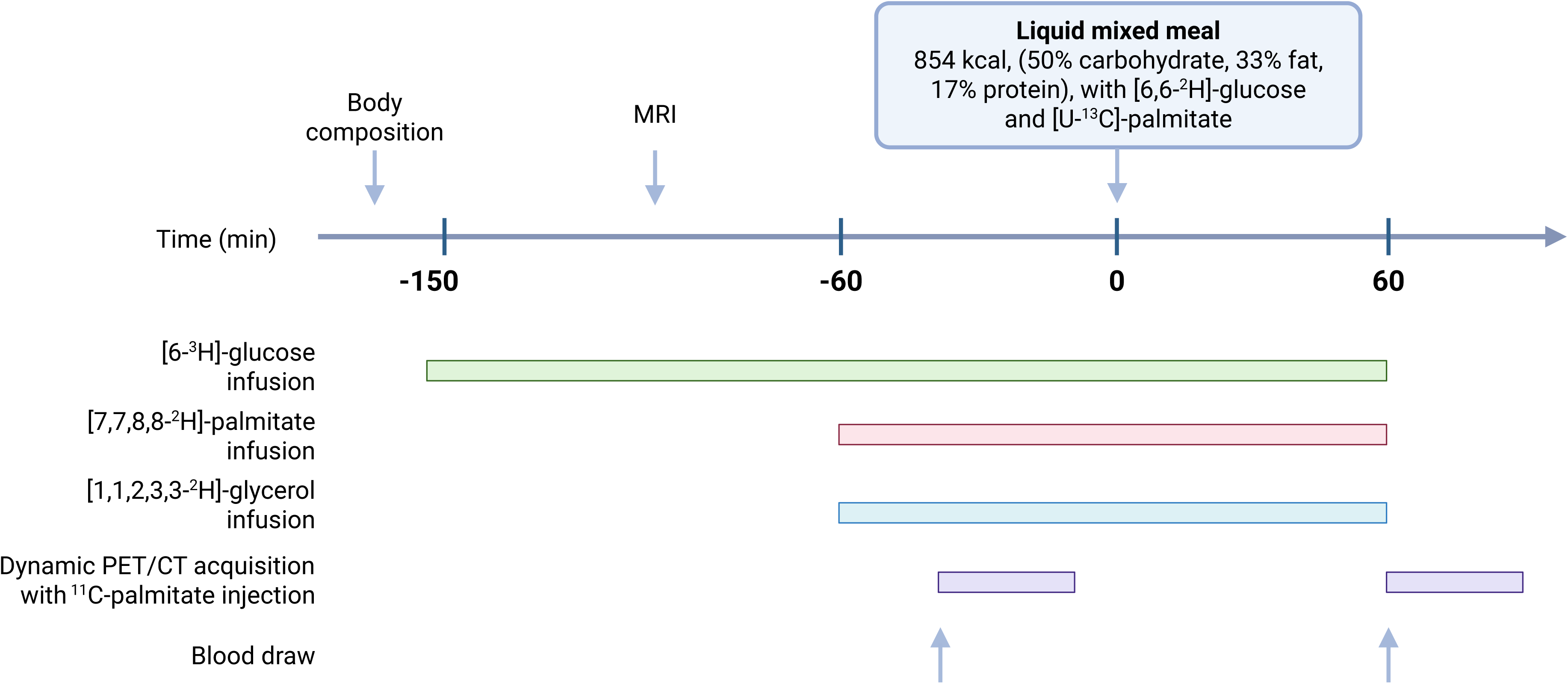
Experimental design of the study. MRI, magnetic resonance imaging

A primed continuous intravenous infusion of [6-^3^H]-glucose was initiated 150 min before meal ingestion and maintained throughout the study (bolus: 1.5 µCi, continuous infusion: 0.25 µCi·min^-1^). Continuous intravenous infusions of [7,7,8,8-^2^H_4_]palmitate (0.1 µmol·kg^-1^·min^-1^, Cambridge Isotope Laboratories), prepared in 25% albumin, and [1,1,2,3,3-^2^H]glycerol (0.010 µmol·kg^-1^·min^-1^, Cambridge Isotope Laboratories) were initiated 60 min before meal ingestion and maintained thereafter^20^. A bolus of sterile [1-^13^C]NaHCO_3_ (1.2 µmol·kg^-1^, Cambridge Isotope Laboratories) was also administrated 60 min before the meal to prime the bicarbonate pool. At time 0, participants ingested a liquid mixed meal over 20 min (500 mL, 849 Kcal, 33% fat, 17% protein and 50% carbohydrates), containing [6,6-^2^H_2_]glucose (0.9 g)^21^ and [U-^13^C]-palmitate (9 µmol·kg^-1^, Cambridge Isotope Laboratories). Blood samples were collected in the fasted state and 60 min after the meal ingestion in tubes containing Na_2_EDTA or 10µL aprotinin·mL^-1^ of blood for the determination of tracer enrichment and circulating metabolite concentrations.

### 2.3 PET and MRI Imaging

CT (30 mAs) and 30-minute dynamic PET acquisition centered on the thoracoabdominal segment were performed using [^11^C]palmitate (Sherbrooke Molecular Imaging Center, CIMS) to assess NEFA fluxes in lean and adipose tissues. PET measurements were performed in the fasted state and repeated 60 min after meal ingestion. [^11^C]palmitate (∼178.3 MBq [SD 35.2]) was administered intravenously over 30 seconds for each acquisition. Images were reconstructed into 35 frames: 1×30 (background frame), 12×5, 6×10, 6×30 and 10×150 seconds. Dynamic and cardiac-gated images were reconstructed from the list-mode acquisition using an iterative algorithm (4 iterations and 5 subsets with gaussian filter of 4 mmm) with point spread function and time of flight modeling. The image matrix was 440 x 440 voxels with an isotropic voxel size of 1.65 mm^3^. Cardiac-gated image acquired 5-20 min post-tracer injection were reconstructed and summed into 16 gates.

MRI was performed in the fasted state for organ-specific fat fraction using a 3T Philips Ingenia scanner (Philips Healthcare). Images were acquired with an 8-channel torso surface coil (dStream; Philips) covering the torso. mDixon Quant images were acquired to quantify the proton density fat fraction (PDFF), with Quant TE=6 echoes, first one 0.97 ms with echo spacing of 0.7 ms, TR=5.67 ms, Flip angle=3°, voxel size=2.08×2.08×6.00 mm^3^.

### 2.4 Image and data Analysis

#### PET/CT

Blood and tissue time-activity curves (TACs) were extracted from dynamic PET images using PMOD quantification software (version 3.7). PET data were corrected for dead time, radioactive decay, photon attenuation, scatter and random coincidences. Volumes of interest were delineated manually and/or semi-automatically on CT images using 3D Slicer^22^ with the TotalSegmentator extension^23^. For manually delineated volumes, a minimum of six contiguous slices was included. For semi-automated segmentations performed with 3D Slicer, the largest possible organ volume was retained. Regions of interest included the left ventricle cavity, liver, myocardium (left ventricle), skeletal muscle (limited to psoas major and paraspinal muscles), visceral abdominal adipose tissue (VAT), and subcutaneous abdominal adipose tissue (SAT). The left ventricular cavity activity was used for the image-derived arterial input function. In addition, Hounsfield units (HU) measured in the left ventricular myocardium on CT images were used as a proxy of myocardial fat content^24^. For ventriculography, the Corridor4DM v5.2 clinical software (Segami; Invia) was used for reorientation and to compute left ventricular function.

Tissue-specific multicompartmental kinetic models of NEFA metabolism were applied to TACs from the heart^25^, liver^26^, skeletal muscle^27^, SAT^27^, and VAT^26^ to estimate fractional NEFA uptake, oxidation, and esterification (**Supplemental Figure 1**). Visual inspection of VAT TACs revealed a rapid early washout phase suggestive of radiorapid tracer tissue clearance (**Supplemental Figure 2**), a pattern that was not observed in SAT TACs and was not adequately captured by the standard adipose tissue model^27^ which does not include an oxidative or metabolic clearance compartment. The multicompartmental kinetic model developed for the liver^26^ was therefore applied to VAT TACs, as it includes an oxidation/metabolic clearance compartment and provided a better representation of VAT tracer kinetics. Model performance was assessed by visual inspection of fitted TACs and residual plots. The selected VAT model adequately fitted the measured TACs, including the rapid early washout phase, with residuals comparable to those obtained using the standard adipose tissue model (**Supplemental Figure 1**). In the liver, the model also allowed estimation of palmitate release into VLDL-TG. In VAT, the slow-release compartment of tracer excretion was interpreted as palmitate secretion from VAT. Because [^11^C]palmitate oxidation releases [^11^C]CO_2_ into the circulation, uncorrected plasma radioactivity may overestimate the [^11^C]palmitate input function. Metabolite correction of the input function was not applied in the main analysis. As circulating [^11^C]CO_2_ was not measured in our participants, we assessed its potential influence by repeating the kinetic analysis in all tissues using data reported by Christensen et al.^28^, applying the values obtained in healthy participants under basal conditions to the fasting scans and those obtained in healthy participants under hyperinsulinemic conditions to the postprandial scans. The direction and significance of changes in fractional and net fluxes were unchanged in all tissues. Results were thus presented without correction. Results on fractional constant rates are displayed in **Supplemental Table 1**. Palmitate uptake represents the sum of palmitate oxidation and esterification. Fractional rates (mL·mL of tissue^-1^·min^-1^) were normalized by 100 g of tissue and thus expressed as mL·100g^-1^·min^-1^ with ρ of 1.06 g·mL^-1^ for the liver, 1.05 g·mL^-1^ for myocardium, 1.04 g·mL^-1^ for skeletal muscle and 0.92 g·mL^-1^ for adipose tissue^29,30^. They were further multiplied by circulating palmitate concentration (assuming that palmitate kinetics are representative of those of the total NEFA pool) to derive tissue-specific net palmitate fluxes (µmol·100g^-1^·min^-1^).

#### MRI

Tissue lipid content was assessed in the liver and skeletal muscle (limited to psoas major and paraspinal muscles). Volumes of interest were segmented on T1-weighted anatomical images using 3D Slicer^22^ with the TotalSegmentator extension^23^ and subsequently applied to the corresponding PDFF maps to extract mean tissue-specific PDFF values expressed as percentages.

For SAT and VAT compartments, fatty acid composition was further characterized by fitting a 10-peak spectral fat model voxel-wise to the complex multi-echo signal using non-linear least squares optimization (trust-region reflective algorithm). This approach simultaneously estimated PDFF, the number of double bonds (NDB), apparent transverse relaxation rate (R2*), and local frequency offset (ΔB₀). Phase convention calibration was performed automatically by selecting the variant minimizing the root mean square error over an adipose tissue reference region. Segmentation masks were morphologically eroded by one iteration to exclude partial volume voxels at tissue boundaries, and only voxels with a PDFF ≥ 20% were retained to minimize contamination from non-adipose tissue. Mean NDB was extracted per compartment for each participant and was used as an index of adipose tissue lipid unsaturation.

### 2.5 Metabolites and hormonal Assays

Total plasma NEFA and triglycerides were measured using specific colorimetric assays (Wako Industrials diagnostics and Thermo DMA, respectively). Insulin was measured by Luminex xMAP-based multiplex immunoassays (Millipore, Billerica, MA). Plasma total glucose and meal [6,6-^2^H_2_]glucose enrichment was measured by liquid chromatography-tandem mass spectrometry (XEVO TQ-S micro Acquity Class I UPLC) and [6-^3^H]glucose activity was determined by liquid scintillation^31^ to calculate endogenous, and exogenous glucose rate of appearance (Ra_glucose_) and disappearance (Rd_glucose_)^32^. [U-^13^C]palmitate, [7,7,8,8-^2^H_4_]palmitate and [1,1,2,3,3-^2^H]glycerol enrichments and levels of palmitate were determined using gas chromatography-tandem mass spectrometry (XEVO TQ-GC Agilent 8890 GC system) to calculate plasma palmitate, NEFA and glycerol appearance rates^8,20^.

### 2.6 Calculations

Plasma rates of appearance of palmitate (Ra_palmitate_), total NEFA (Ra_NEFA_) and glycerol (Ra_glycerol_) were calculated using the Steele non-steady-state equations, as previously described^15^. Rates of appearance of endogenous glucose (Ra_endo glucose_) and exogenous glucose from the meal (Ra_exo glucose_), as well as glucose rate of disappearance Rd_glucose_ were calculated as previously described^32^. Postprandial changes in organ-specific NEFA fluxes, metabolites concentrations, and rates of appearance and disappearance were calculated as the difference between postprandial (60min after meal ingestion) and fasting values.

Hepatic insulin resistance was estimated as the product of fasting endogenous glucose production (Ra_glucose_) and fasting insulin concentration, and adipose tissue insulin resistance was estimated as the product of fasting Ra_NEFA_ and fasting insulin concentration. Body mass, FM and LM were divided by height squared to derive body mass index (BMI), fat mass index (FMI) and lean mass index (LMI), respectively.

### 2.7 Statistical Analyses

Variables reported for descriptive purposes only are presented as mean (95%CI) or median [interquartile range, IQR]. Linear mixed-effects models were used to examine the effect of fasting versus postprandial state (fixed effect), on circulating insulin, glycerol, glucose and fatty acid kinetics, and organ-specific fatty acid fluxes. A subject-specific random intercept was incorporated to account for repeated measurements and inter-individual variability. Different covariance structures were tested and selected according to Akaike and Bayesian information criteria.

Statistical inference focused on estimating the within-subject change from fasting to the postprandial state. Therefore, all model contrasts were defined as postprandial minus fasting. Positive estimated differences indicate higher values in the postprandial state than in the fasting state, whereas negative estimated differences indicate lower postprandial values.

Unless otherwise specified, model-derived results are presented as least-square means (LSmeans) with 95% confidence interval (95%CI) and fasting-to-postprandial changes are presented as estimated mean differences with their 95%CI. For all models, residual normality was examined using Pearson residuals, residuals-versus-fitted plots, QQ plots, residual distributions by nutritional state, and Shapiro-Wilk tests. When necessary, dependent variables were log-transformed. For log-transformed outcomes, back-transformed LSmeans (geometric mean) with 95% CI are reported and fasting-to-postprandial effects are expressed as percentage change, derived from the exponentiated contrast of the linear mixed-model on the log scale, with 95% CI obtained by back-transforming the corresponding confidence limits. Positive percentage changes indicate higher estimated postprandial values compared with fasting values, whereas negative percentage changes indicate lower estimated postprandial values.

A defective batch of [7,7,8,8-^2^H_4_]palmitate produced implausible plasma enrichments in a subset of participants. These data were identified from the enrichment curves and were excluded. Valid data was available in 22 participants for the mixed-model analyses, which include participants with a valid value at one of the two time points. Correlation analyses require a fasting-to-postprandial change and were therefore restricted to the 20 participants with a valid value at both time points.

Exploratory associations between within-subject fasting-to-postprandial changes in tissue-specific fatty acid fluxes or model-derived kinetic parameters and selected metabolic, anthropometric, hemodynamic, and imaging variables were assessed using Spearman rank correlations. Individual changes were calculated as postprandial minus fasting values. Organ-specific fatty acid fluxes and kinetic parameters were considered as outcome variables, whereas circulating metabolites, NEFA Ra and glucose Ra and Rd, insulin resistance indices, body composition indices, tissue fat fraction or Hounsfield units, and hemodynamic variables when applicable were considered as candidate correlates. Correlation analyses were restricted to predefined biologically relevant pairs. To account for multiple testing, P values were adjusted using the Benjamini-Hochberg false discovery rate procedure separately for each tissue-specific fatty acid flux or kinetic parameter across the candidate variables tested for that outcome. Exploratory correlations were considered statistically significant at q<0.05. Nominal associations with P<0.05 but q≥0.05 were reported descriptively.

All statistical analyses were performed in R (version 4.4.2). Statistical significance for mixed-model analyses was set at P <0.05. Figures were created with Prism 11 (GraphPad, San Diego, CA, USA).

## 3 Results

### 3.1 Participant characteristics

Participant characteristics are presented by sex in **Table 1**. Overall, participants had a mean age of 59 years (95%CI 57 to 62), body mass of 87.6 kg (76.4 to 98.8) and BMI of 31.8 kg·m^-2^ (29.9 to 33.7). Of the 24 women, 19 were postmenopausal. Waist circumference averaged 97 cm (93 to 102) in women and 108 cm (101 to 114) in men. Mean lean mass was 40.1 kg (37.7 to 42.5) in women and 57.4 kg (53.0 to 61.8) in men, and fat mass was 41.3 kg (35.9 to 46.7) in woman and 36.1 kg (28.6 to 43.6) in men. Median liver fat fraction was 9.0 % [Q1-Q3 6.0-20.2] overall, with highest values in men compared to women (10.9 % [7.6 to 18.8] vs 8.6 % [5.9 to 22.7]).

### 3.2 Meal ingestion increased glucose disposal and cardiac output while suppressing adipose tissue lipolysis

Meal ingestion induced marked changes in circulating metabolites and whole-body substrate kinetics (**Table 2**). Plasma insulin increased by 548 % (95%CI 416 to 716) (P<0.0001) in the 60 min after meal ingestion, together with a significant rise in plasma glucose (+3.1 mmol·L^-1^ [95% CI 2.2 to 3.9], P<0.0001). Consistent with these postprandial changes, Ra_endo glucose_ decreased (−782 µmol·min^-1^ [-1103 to −461], P<0.0001) 60min after meal ingestion, whereas Rd_glucose_ increased (+1438 µmol·min^-1^ [1112 to 1765], P<0.0001). Ra_exo glucose_ reached 2213 µmol·min^-1^ [1827 to 2598] in the postprandial state. Plasma NEFA and palmitate concentrations decreased markedly after meal ingestion (−484 µmol·L^-1^ [-581 to −387], P<0.0001; −107 µmol·L^-1^ [-121 to −92], P<0.0001 respectively), accompanied by a significant reduction in whole-body Ra_NEFA_ (−400 µmol·min^-1^ [-624 to −176], P=0.001), consistent with suppression of adipose tissue lipolysis. Although plasma glycerol concentration decreased in postprandial state (−37 % [-44 to −30], P<0.0001), the decrease in Ra_glycerol_ was statistically not significant (−10% [-26 to 10], P=0.298), likely reflecting an increased contribution from the intravascular lipolysis of triglyceride-rich lipoprotein. Cardiac output increased significantly in the postprandial state (+643 mL·min^-1^ [355 to 932], P=0.0001; **Table 2**).

**Table 2:**
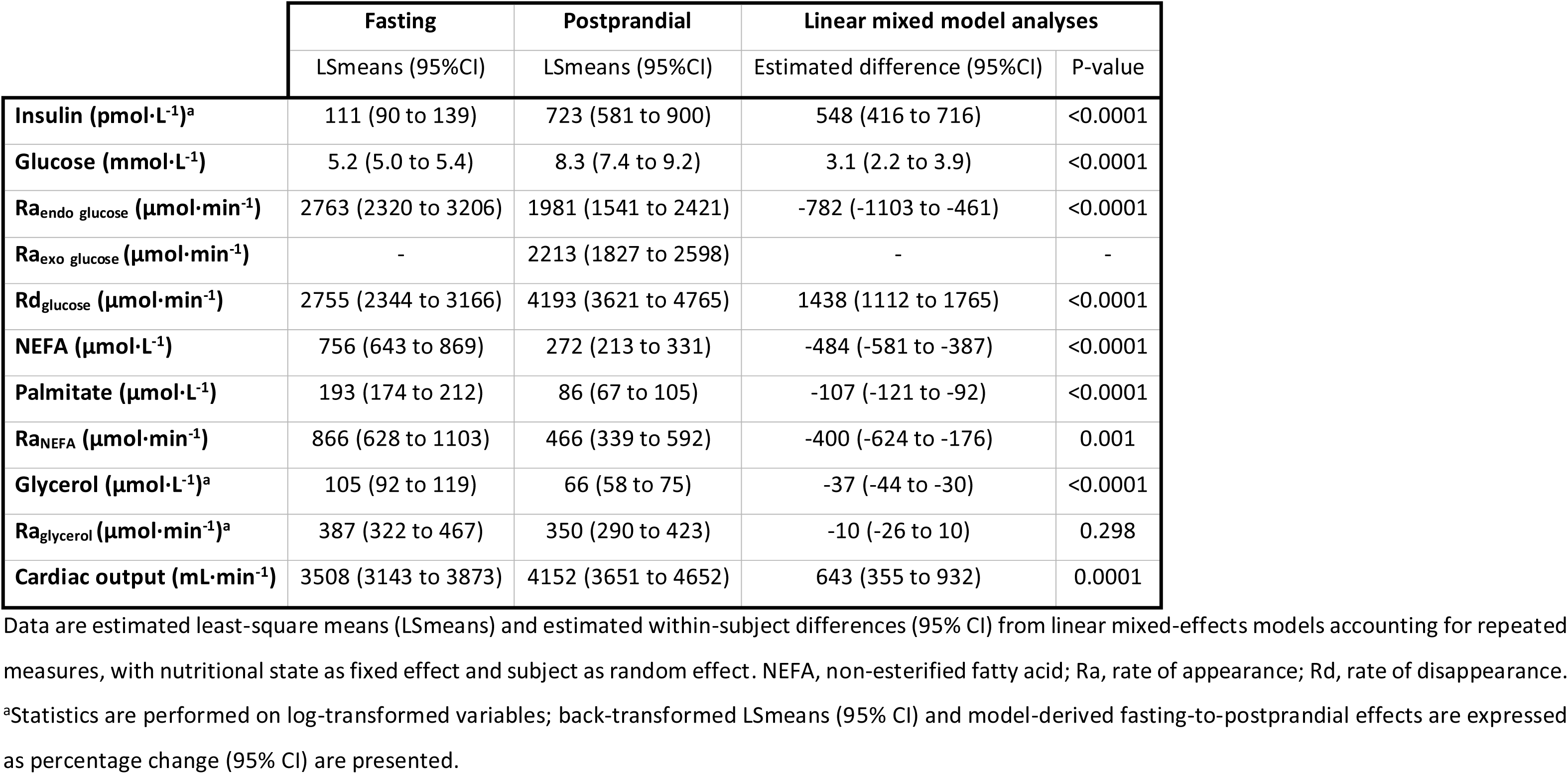
Circulating metabolites, whole-body substrate kinetics, and cardiac output in the fasting and postprandial states.

### 3.3 Meal ingestion reduces net palmitate uptake in both adipose depots, increases fractional esterification, and suppresses palmitate release from VAT

In VAT, net palmitate uptake (−56 % [-67 to −41], P<0.00001; **Figure 2A left**), esterification (−50 % [-57 to −42], P<0.0001; **Figure 2B left**), oxidation (−64 % [-79 to −38], P<0.0001; **Figure 2C left**) and release (−100 % [-100 to −91], P=0.001; **Figure 2D left**) decreased from fasting to postprandial conditions. Fractional esterification rate increased (+0.51 ml·100g^-1^·min^-1^ [0.12 to 0.89], P=0.012; **Figure 2B right**) and fractional palmitate release rate decreased (−99 % [-100 to −77], P=0.005; **Figure 2D right**), whereas fractional palmitate uptake rate (+7 % [-16 to 36], P=0.565; **Figure 2A right**) and fractional oxidation rate (+2.2 ml·100g^-1^·min^-1^ [-2.5 to 6.9], P=0.345; **Figure 2C right**) did not differ.

**Figure 2:**
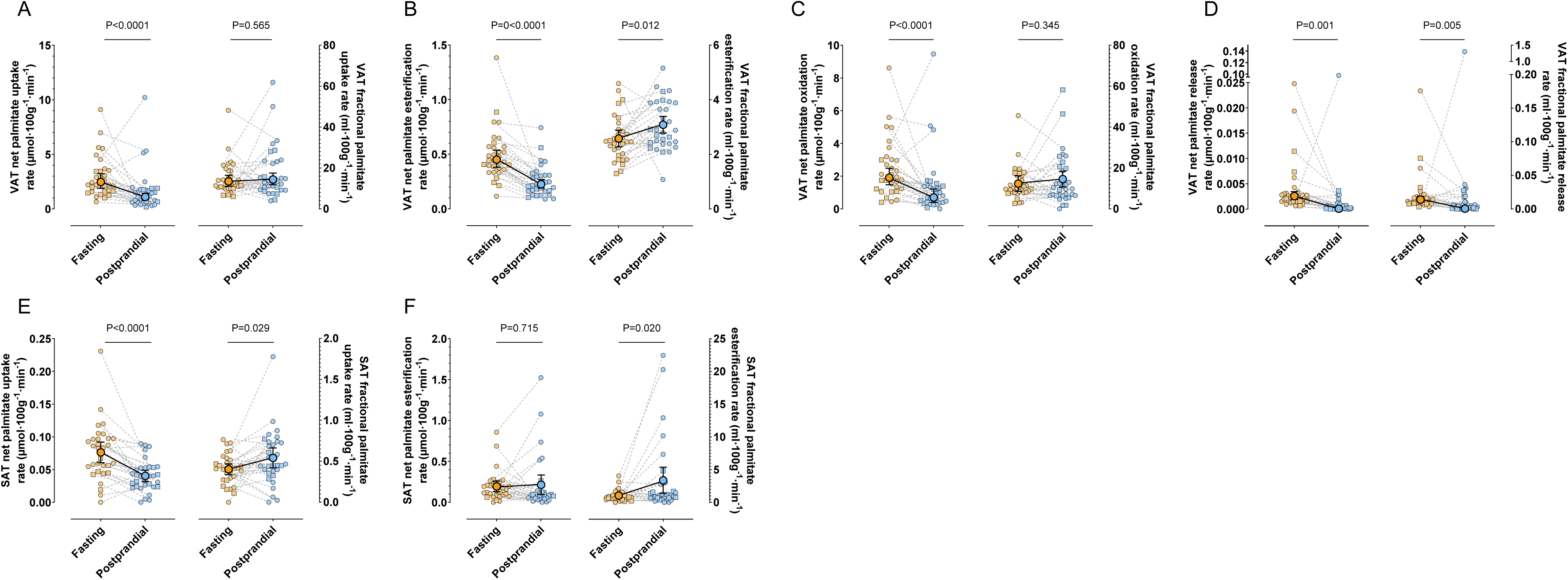
Fractional palmitate esterification increases in both adipose tissues and palmitate release decreased in VAT after a meal. Visceral adipose tissue (VAT) palmitate uptake (A), esterification (B), and oxidation (C), palmitate release (D) as well as subcutaneous adipose tissue (SAT) palmitate uptake (E) and esterification (F), are presented as LSmeans with 95% confidence interval (CI) from mixed-effects models accounting for repeated measures, with nutritional state (fasting vs postprandial) included as a fixed effect; for VAT net uptake, esterification, and net and fractional palmitate release, models were fitted on log-transformed data and back-transformed LSmeans and 95%CI are presented. Left panels show net palmitate flux rates, calculated as the product of fractional rates and circulating palmitate concentration, whereas right panels show fractional rates. Palmitate uptake is the sum of palmitate esterification and oxidation. Individual data points are shown as circles for women and squares for men.

In SAT, net palmitate uptake decreased (−0.04 µmol·100g^-1^·min^-1^ [-0.05 to −0.02], P<0.0001; **Figure 2E left**), whereas net esterification did not differ (+0.02 µmol·100g^-1^·min^-1^ [-0.09 to 0.13], P=0.715; **Figure 2F left**) between fasting and postprandial conditions. Fractional palmitate uptake increased (+0.14 ml·100g^-1^·min^-1^ [0.02 to 0.26], P=0.029; **Figure 2E right**), together with a higher fractional esterification rate (+2.3 [0.4 to 4.3] ml·100g^-1^·min^-1^, P=0.020; **Figure 2F right**).

### 3.4 Lower postprandial palmitate availability reduces net uptake while lean tissue extraction increases, with divergent intracellular metabolic fates

In the heart, net palmitate uptake decreased from fasting to postprandial conditions (−43 % [-50 to −34], P<0.0001; **Figure 3A left**), whereas net esterification also decreased (−76 % [-83 to −65], P<0.0001; **Figure 3B left**), and net oxidation did not differ (+0.1 µmol·100g^-1^·min^-1^ [-0.5 to 0.8], P=0.736; **Figure 3C left**) postprandially. Fractional fluxes showed a different pattern after the meal, with a lower fractional esterification rate (−42 % [-60 to −17], P=0.005; **Figure 3B right**) and a higher fractional oxidation rate (+12.8 mL·100g^-1^·min^-1^ [9.4 to 16.3], P<0.0001; **Figure 3C right**), together with a higher fractional palmitate uptake rate (+8.1 mL·100g^-1^·min^-1^ [5.4 to 10.9], P<0.0001; **Figure 3A right**).

**Figure 3:**
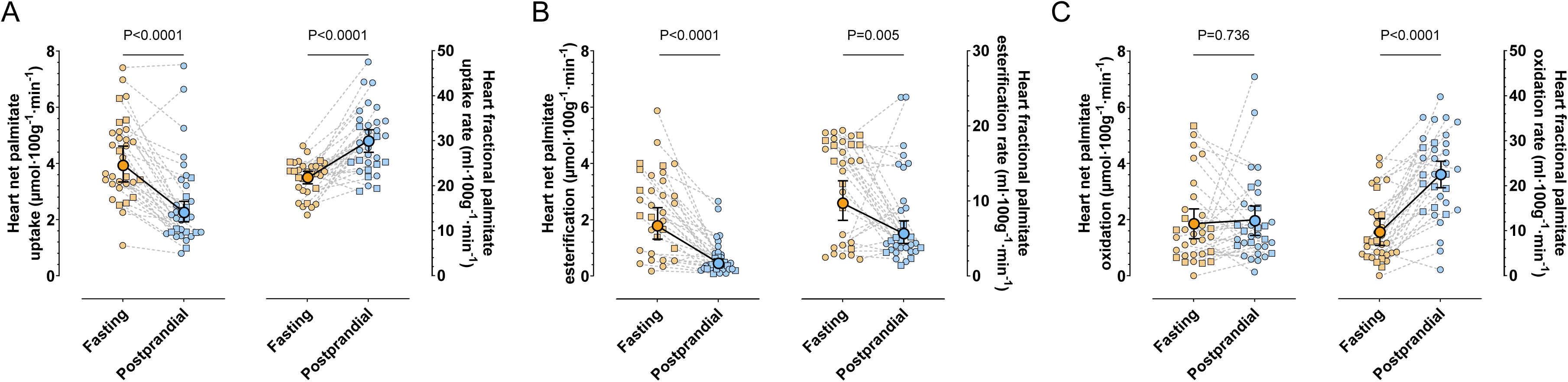
Fractional palmitate uptake and oxidation increase in the heart after a meal, while fractional esterification decreases. Cardiac palmitate uptake (A), esterification (B) and oxidation (C) are LSmeans with 95% confidence interval (CI) from mixed-effects models accounting for repeated measures, with nutritional state (fasting vs postprandial) as fixed effect; for net uptake, esterification and fractional esterification, models were fitted on log-transformed data and back-transformed LSmeans and 95%CI are presented. Left panels show net palmitate flux rates, computed as the product of fractional rates and circulating palmitate concentration, and right panels represent fractional rates. Palmitate uptake is the sum of palmitate esterification and oxidation. Individual data points are shown as circles for women and squares for men.

In skeletal muscle, net palmitate uptake (−48 % [-57 to −37], P<0.0001; **Figure 4A left**), esterification (−0.03 µmol13 mL·100g^-1^·min^-1^ [-0.05 to −0.02], P<0.001; **Figure 4B left**), and oxidation (−47 % [-59 to −32], P<0.0001; **Figure 4C left**) decreased from fasting to postprandial conditions. Fractional palmitate uptake increased slightly (+20 % [2 to 41], P=0.030; **Figure 4A right**), whereas fractional palmitate esterification (+0.10 mL·100g^-1^·min^-1^ [-0.04 to 0.24], P=0.140; **Figure 4B right**) and oxidation (+21 % [-4 to 53], P=0.095; **Figure 4C right**) rates were unchanged postprandially.

**Figure 4:**
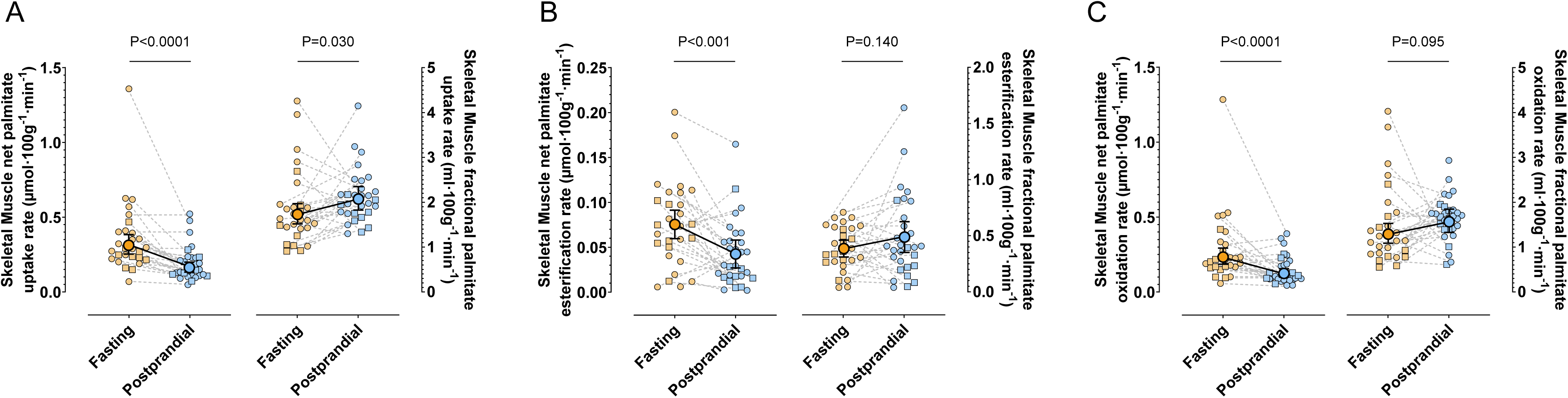
Fractional palmitate uptake increases in skeletal muscle after a meal, without changes in esterification, oxidation. Skeletal muscle palmitate uptake (A), esterification (B) and oxidation (C) are LSmeans with 95% confidence interval (CI) from mixed-effects models accounting for repeated measures, with nutritional state (fasting vs postprandial) as fixed effect; for uptake and oxidation, models were fitted on log-transformed data and back-transformed LSmeans and 95%CI are presented. Left panels show net palmitate flux rates, computed as the product of fractional rates and circulating palmitate concentration, and right panels represent fractional rates. Palmitate uptake is the sum of palmitate esterification and oxidation. Individual data points are shown as circles for women and squares for men.

In the liver, net palmitate uptake (−2.7 µmol·100g^-1^·min^-1^ [-3.2 to −2.2], P<0.0001; **Figure 5A left**), esterification (−43 % [-49 to −36], P<0.0001; **Figure 5B left**), and oxidation (−1.1 µmol·100g^-1^·min^-1^ [-1.5 to −0.6], P<0.0001; **Figure 5C left**) were reduced after meal intake, and net palmitate release into VLDL did not differ (−20 % [-49 to 25], P=0.313; **Figure 5D left**). In contrast, fractional palmitate uptake rate was higher postprandially (+29 % [21 to 39], P<0.0001; **Figure 5A right**), mainly driven by a higher fractional esterification rate (+7.3 mL·100g^-1^·min^-1^ [5.3 to 9.4], P<0.0001; **Figure 5B right**), while fractional oxidation rate remained unchanged (+2.8 mL·100g^-1^·min^-1^ [-0.6 to 6.2], P=0.101; **Figure 5C right**). Fractional VLDL-palmitate release was also higher in response to meal ingestion (+90 % [20 to 200], P=0.007; **Figure 5D right**) as compared to fasting conditions.

**Figure 5:**
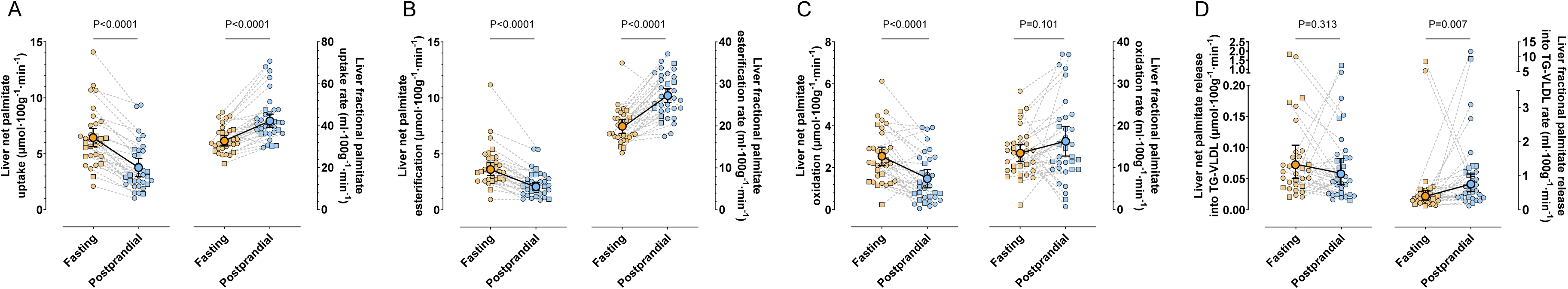
Fractional palmitate uptake, esterification, and palmitate release into VLDL increase in the liver after a meal. Liver palmitate uptake (A), esterification (B), oxidation (C), and palmitate release into VLDL (D) are presented as LSmeans with 95% confidence interval (CI) from mixed-effects models accounting for repeated measures, with nutritional state (fasting vs postprandial) included as a fixed effect; for net palmitate esterification and VLDL-palmitate release and fractional palmitate uptake and VLDL-palmitate release models were fitted on log-transformed data and back-transformed LSmeans and 95%CI are presented. Left panels show net palmitate flux rates, calculated as the product of fractional rates and circulating palmitate concentration, whereas right panels show fractional rates. Palmitate uptake represents the sum of palmitate esterification and oxidation. Individual data points are shown as circles for women and squares for men

### 3.5 Exploratory associations between fasting-to-postprandial changes in tissue fatty acid fluxes and metabolic or body composition variables

Exploratory Spearman rank correlations were used to test whether the fasting-to-postprandial changes in tissue-specific fractional palmitate fluxes were associated with changes in circulating insulin concentrations, whole-body glucose and NEFA kinetics, anthropometric measures, or organ-specific lipid content All associations are shown in **Figure 6** with associations surviving FDR corrections highlighted by a black square.

**Figure 6:**
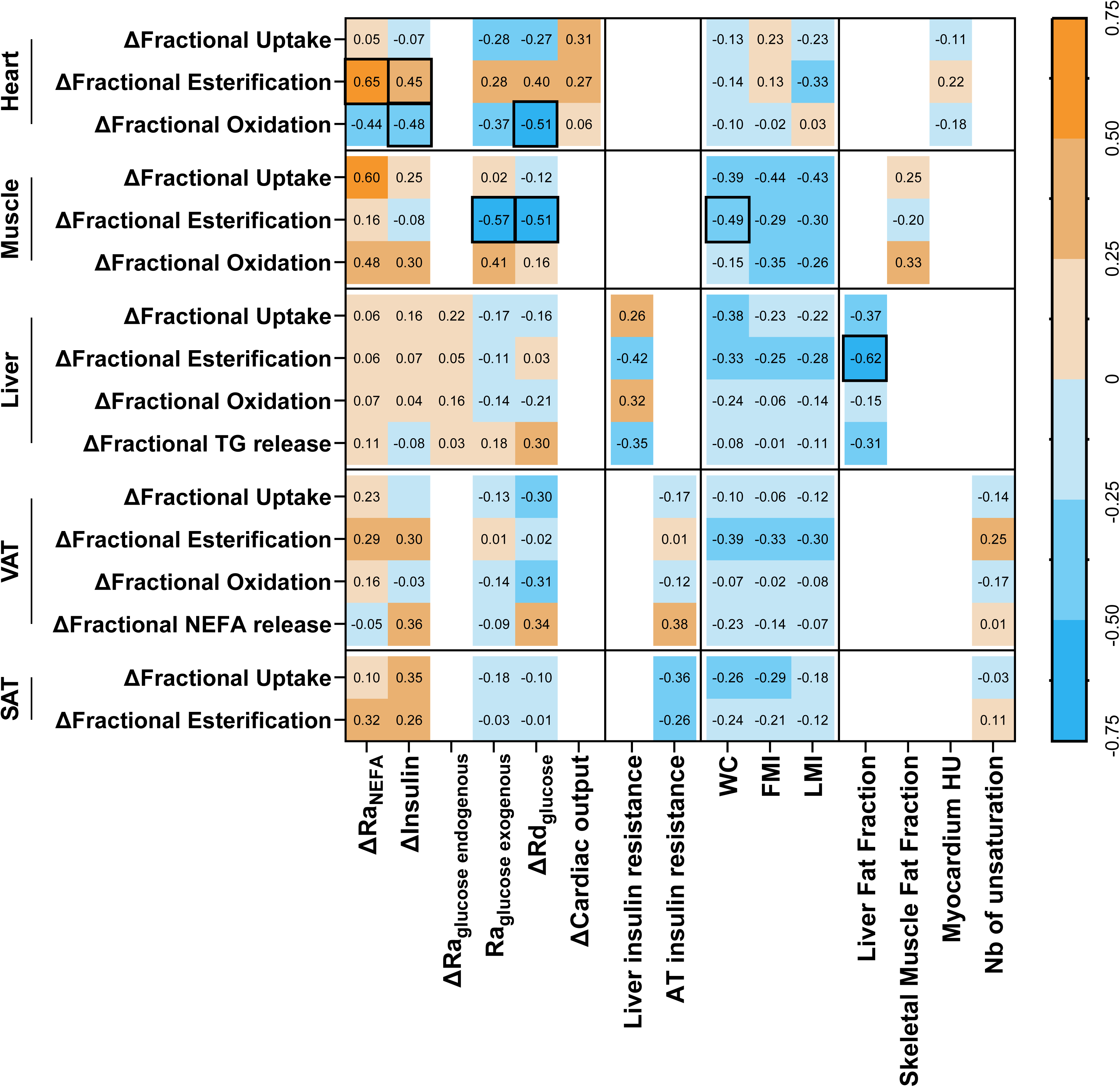
Exploratory associations between fasting-to-postprandial changes in fractional palmitate fluxes and metabolic, anthropometric, and imaging determinants. Spearman rank correlations were used to assess associations between within-subject fasting-to-postprandial changes in tissue-specific fractional palmitate fluxes and predefined potential determinants, including circulating metabolites, whole-body NEFA and glucose kinetics, insulin resistance indices, body composition indices, and tissue fat content or attenuation measures. Individual changes were calculated as postprandial minus fasting values. Correlation analyses were restricted to predefined biologically relevant pairs. Cells show Spearman correlation coefficients (rho). P-values were adjusted for multiple testing using the Benjamini-Hochberg false discovery rate procedure separately for each flux variable. Correlations considered statistically significant after FDR correction are highlighted by a black outline around the rho value.

In VAT and SAT, no associations between the changes in fractional palmitate fluxes and metabolic, body composition variables or adipose tissue unsaturation level survived FDR correction.

In the heart, the change in fractional palmitate esterification was positively correlated with the changes in Ra_NEFA_ (rho=0.65, q=0.046, n=16) and with insulin concentrations (rho=0.45, q=0.046, n=31), and the change in fractional NEFA oxidation was negatively correlated with the changes in Rd_glucose_ (rho=-0.51, q=0.031, n=27) and with insulin concentrations (rho=-0.48, q=0.031, n=31). Thus, participants with the smallest rise in insulin had the largest decrease in cardiac fractional palmitate esterification and the largest increase in cardiac fractional palmitate oxidation. The change in cardiac fractional palmitate esterification was also positively associated with Rd_glucose_ changes but only nominally (rho=0.40, P=0.039).

In skeletal muscle, the change in fractional palmitate esterification was negatively correlated with the postprandial Ra_exo glucose_ (rho=-0.57, q=0.042, n=21), with the change in Rd_glucose_ (rho=-0.51, q=0.042, n=22), and with waist circumference (rho=-0.49, q=0.042, n=26). Thus, participants with a higher postprandial Ra_exo glucose_, a larger increase in Rd_glucose_, or a larger waist circumference had a lower skeletal muscle fractional palmitate esterification after the meal relative to fasting. The change in fractional palmitate uptake was positively correlated with the change in Ra_NEFA_ (rho=0.60, P=0.031), but only nominally.

In the liver, the change in fractional palmitate esterification was negatively correlated with liver fat content (rho=-0.62, q=0.006, n=27). Thus, participants with a higher liver fat had a smaller postprandial increase in hepatic fractional palmitate esterification after the meal. The change in hepatic fractional palmitate esterification was also negatively correlated with the liver insulin resistance index, but only nominally (rho=-0.42, P=0.027).

## 4 Discussion

In this study, we combined dynamic [^11^C]palmitate PET/CT with stable isotope tracers and MRI to characterize the fasting-to-early-postprandial changes in tissue-specific NEFA uptake and intracellular fate across lean organs and adipose tissues in individuals with prediabetes. As circulating NEFA fell after the meal, net NEFA uptake decreased in all tissues. It shifted toward oxidation in the heart and toward esterification in the liver and both adipose depots, but did not change in skeletal muscle. As a result, net oxidation was maintained in the heart and net VLDL-palmitate release was maintained in the liver, while NEFA release from VAT was almost completely suppressed. VAT also showed a large palmitate oxidative rate, which has not been described previously. Thus, the postprandial fall in plasma NEFA concentrations does not translate into a proportional reduction in NEFA delivery to tissues.

It has been shown previously that at the whole-body level, lower Ra_NEFA_ is associated with greater NEFA clearance in healthy men^6^, suggesting an inverse relationship between NEFA availability and disposal. Accordingly, the marked suppression of circulating fatty acids after an oral glucose load can increase whole-body palmitate clearance by more than twofold^33^. Our tissue-level findings are consistent with this relationship. However, the change in systemic Ra_NEFA_ was not associated with the changes in fractional uptake in any tissue after correction for multiple testing. In skeletal muscle, a smaller fall in Ra_NEFA_ was nominally associated with a larger increase in fractional uptake, which is opposite to the expected inverse relationship. This suggests that the increase in tissue extraction cannot be fully explained by changes in NEFA availability. Similarly, although insulin is a major regulator of whole-body NEFA clearance through suppression of adipose tissue lipolysis^6^, the postprandial increase in insulin was not associated with the change in fractional NEFA uptake in any tissue and was associated with the intracellular fate of NEFA only in the heart, as discussed below. Whether the absence of association with uptake reflects reduced insulin sensitivity of these tissues in prediabetes or limited statistical power cannot be determined without a metabolically healthy comparison group.

In both adipose depots, net palmitate uptake decreased after the meal. In VAT, net palmitate esterification and oxidation also decreased, and palmitate release was almost completely suppressed. However, fractional palmitate esterification increased in both depots. In VAT, fractional palmitate uptake did not change, and a larger share of the NEFA taken up was esterified. The increase in fractional palmitate esterification is consistent with the established action of insulin on white adipose tissue, which suppresses intracellular triglyceride lipolysis and promotes fatty acid uptake and esterification^4,5,14^. In healthy individuals, fractional re-esterification of fatty acids roughly doubles across the physiological range of insulin concentrations^5^, and direct in vivo measurements in SAT show a rise from approximately 20% in the fasted state toward near-complete re-esterification during hyperinsulinemia^7^. Our measurements concern palmitate taken up from the circulation rather than palmitate released within the adipocyte from triglyceride lipolysis, but esterification of fatty acids from both sources are promoted by insulin. Yet, in our participants, the increase in fractional palmitate esterification in adipose tissues was not associated with the magnitude of the insulin response.

The reduction in VAT NEFA release reflects lower rate of recycling of newly taken-up fatty acids rather than reduced lipolysis of pre-existing triglyceride stores. [^11^C]palmitate-PET tracks labelled fatty acids that entered the adipocytes, were esterified, and were then released. Thus, after the meal, VAT retained a larger share of the fatty acids it took up instead of releasing them back into the circulation. Because VAT drains into the portal vein, this reduces NEFA delivery to the liver. Moreover, individuals with more intra-abdominal fat, but not more subcutaneous fat, show a smaller suppression of plasma NEFA by insulin^34^.

Importantly, VAT showed a pattern of NEFA kinetics not previously described. Unlike SAT, VAT time-activity curves showed a rapid early washout of [^11^C]palmitate (**Supplemental Figure 2**) that the standard adipose tissue model could not describe. The latter model was developed in SAT^27^, the depot most often studied. Since VAT can only be sampled during surgery, and because it drains into the portal vein, its fatty acid fluxes can only be estimated indirectly in humans. VAT kinetics required a model with an oxidation (or metabolic clearance) compartment, and oxidation accounted for a median of 82 % [Q1-Q3 79-87] of palmitate uptake by VAT in the fasted state, and 77 % [68-85] after the meal. To our knowledge, such a large oxidative component has not been described in human VAT. A higher oxidative component is consistent with the higher mitochondrial content and mass-specific respiration rate in VAT, compared to SAT^35^. VAT is also a highly dynamic adipose depot, rapidly changing in response to energy excess or deficits. Weight loss interventions, including pharmacotherapies and bariatric surgery, reduce VAT proportionally more than SAT^36^, and GLP-1 receptor agonists reduce VAT and SAT areas by similar absolute amounts despite the smaller volume of VAT^37^. However, how VAT handles NEFA in the short term, after an overnight fast or after a meal, was unknown. Our results show that VAT NEFA metabolism also changes rapidly after a meal.

In the heart, fractional palmitate uptake increased after the meal ingestion, together with a shift from esterification toward oxidation. This occurred while glucose and insulin rose and circulating NEFA fell, conditions that classically favor a shift away from fatty acid oxidation^38^. Cardiac output also rose after the meal, and an acute increase in cardiac workload can increase myocardial fatty acid uptake and oxidation^39^, fatty acids being the main source of ATP in the heart^40^. The postprandial rise in cardiac NEFA oxidation could also reflect a reduced capacity of the heart to switch between substrates in prediabetes, or the increase in cardiac workload discussed below. Without a metabolically healthy comparison group, however, reduced capacity to switch substrates cannot be established. Insulin also promotes the translocation of CD36 to the sarcolemma, although most evidence to date is limited to *ex vivo* experiments^39,41^. In humans, myocardial fatty acid uptake measured by [^11^C]palmitate is reduced in carriers of a CD36 null mutation, including after an OGTT^33^. A larger postprandial increase in insulin was associated with a smaller decrease in cardiac fractional palmitate esterification and a smaller increase in fractional palmitate oxidation after the meal. Thus, participants with a smaller rise in insulin and glucose relied more on fatty acid oxidation, which is compatible with competition between glucose and fatty acid oxidation. In addition, a smaller fall in systemic Ra_NEFA_ was associated with a smaller decrease in fractional esterification. As the changes in insulin were not correlated with Ra_NEFA_ (rho=0.30, P=0.175), these associations may reflect distinct components of the postprandial response. Whether a greater reliance on NEFA oxidation after a meal contributes to the development of diabetic cardiomyopathy or later cardiac dysfunction in individuals with prediabetes remains unknown and warrants further investigation.

In skeletal muscle, net palmitate uptake, esterification and oxidation decreased after the meal. Fractional palmitate uptake increased modestly, without a significant change in fractional esterification or oxidation. Participants with a higher postprandial appearance of meal-derived glucose or a greater increase in glucose disposal had a lower fractional palmitate esterification rate after the meal relative to fasting. This is opposite to what substrate competition would predict, as greater glucose and insulin availability normally favors fatty acid esterification over oxidation. Whether this reflects altered NEFA handling by muscle in prediabetes cannot be determined without a metabolically healthy comparison group. Fractional esterification was also lower in participants with a larger waist circumference. These exploratory associations should be confirmed in larger studies.

In the liver, fractional palmitate uptake increased, driven by a higher fractional palmitate esterification, while fractional palmitate oxidation did not change. To our knowledge, postprandial hepatic NEFA fluxes have not been measured with this approach in humans. In pigs under an insulin clamp, the same [^11^C]palmitate kinetic model showed no change in the fractional uptake rate, so that net uptake fell in proportion to the reduction in circulating fatty acids^26^. This suggests that systemic hyperinsulinemia alone does not explain the increase in hepatic extraction observed after the meal. An acute increase in hepatocyte transport capacity is also unlikely, as FATP2 and FATP5 are not known to undergo insulin-dependent translocation^42,43^, and hepatic uptake in response to OGTT is preserved in humans lacking CD36^33^. Unlike a peripheral insulin clamp, a meal delivers insulin and glucose to the liver through the portal vein. A meal also increases hepatic blood flow^44^, and both could contribute to the rise in fractional NEFA uptake.

In humans in the fasted state, hepatic fatty acid esterification is mainly determined by systemic and portal fatty acid appearance^26^. After the meal, although net palmitate esterification decreased with lower NEFA availability from circulation, fractional palmitate esterification increased. This suggests that substrate availability alone does not determine postprandial hepatic metabolic partitioning. Participants with more liver fat content showed a smaller postprandial increase in fractional palmitate esterification. In post hoc analyses, liver fat content was also negatively correlated with fasting fractional palmitate esterification (rho=-0.39, P=0.043), but not with fasting net palmitate esterification (rho=-0.14, P=0.489). Hepatic insulin resistance associated with steatosis could contribute to the smaller postprandial increase in hepatic fractional palmitate esterification. Consistent with this, the change in fractional palmitate esterification was also negatively associated with the liver insulin resistance index (rho=-0.42, P=0.027).

Fractional VLDL-palmitate release doubled after the meal, consistent with the postprandial increase in fractional palmitate esterification in the liver, while net VLDL-palmitate release was unchanged. As fatty acid availability is a major determinant of VLDL-TG production^45^ and insulin directly suppresses VLDL particle secretion even when fatty acid availability is maintained, the lower NEFA availability and higher insulin after the meal would be expected to reduce VLDL-TG release^45^. In healthy lean men, a test meal suppresses VLDL-TG secretion by about a half, whereas this suppression is absent in men with obesity and type 2 diabetes^46^. Similarly, insulin acutely suppresses VLDL particle secretion in individuals with less than 5.5% liver fat, but not in those with higher liver fat^34^. Most of our participants had a liver fat content above this threshold. Prediabetes and hepatic steatosis could therefore explain the absence of postprandial suppression of net VLDL-palmitate release in the present study.

### Strength and limitations

This study used state-of-the-art methods to address in a comprehensive way the fasting to postprandial changes in systemic and organ-specific NEFA metabolism. NEFA uptake and intracellular fate were measured simultaneously in five tissues of the same individuals and combined with whole-body glucose and NEFA kinetics and MRI-based measures of organ fat. Dynamic PET with compartmental modeling allowed net and fractional fluxes to be distinguished, which was essential to interpret the postprandial response. Net fluxes depend on circulating palmitate concentrations and decreased in all tissues after the meal. Fractional palmitate fluxes reflect the capacity of each tissue to extract and metabolize NEFA from plasma, independently of their concentration. Beyond their physiological interest, our findings have clinical implications. Because lean tissues increased their fractional palmitate uptake postprandially, plasma NEFA concentrations may underestimate lean tissue exposure to fatty acids in individuals with prediabetes after meals, a state in which humans spend most of the day. This could contribute to ectopic lipid accumulation and the progression toward type 2 diabetes^47,48^. Identifying tissue-specific responses at an early stage of the disease, such as the smaller hepatic response in individuals with more liver fat and the greater reliance of the heart on fatty acid oxidation in those with a weaker insulin response, may point to interventions aimed at preventing complications associated with type 2 diabetes such as MASLD and cardiac failure.

This study has also limitations. Measurements were made at a single time point, 60 min after the meal, so the time course of the responses over the full postprandial period remains unknown. All participants had prediabetes, and a metabolically healthy comparison group would be needed to establish which findings are specific to this population. The data come from a secondary analysis of a trial that was not designed for these outcomes, and the cohort included more women than men, so sex differences could not be examined as a primary analysis. [^11^C]palmitate is a saturated fatty acid tracer, and the intracellular oxidation vs. esterification of oleate (a monounsaturated fatty acid, which make up most of the circulating NEFA pool) and linoleate and other polyunsaturated fatty acids may differ. Net fluxes also assume that palmitate kinetics are representative of the total NEFA pool. Because tissue perfusion was not measured, a contribution of substrate delivery to the intrinsic changes in transport and intracellular metabolism cannot be fully distinguished but unlikely impacts intracellular fate. Finally, the exploratory correlations involved many tests in a modest sample, and the associations that did not survive correction for multiple testing should be regarded as hypothesis-generating.

## 5 Conclusion

In individuals with prediabetes, fractional palmitate uptake increases in the heart, the liver, SAT, and skeletal muscle postprandially. Thus, exposure of lean tissues to circulating palmitate is higher than what could be predicted from the decline of systemic palmitate level and appearance rate after a meal. Furthermore, the intracellular fate of palmitate differed markedly between tissues. In the heart, palmitate shifted from esterification toward oxidation, so that net postprandial palmitate oxidation was maintained. In the liver, palmitate shifted toward esterification, and net VLDL-palmitate release was maintained. In both adipose depots, fractional palmitate esterification increased, and palmitate release from VAT was almost completely suppressed. Also, a large fraction of palmitate taken up by VAT appeared to be oxidized, in contrast to SAT. Postprandial insulin excursion partly predicted these responses in the heart, whereas glucose disposal and waist circumference partly predicted that in skeletal muscle, and liver fat content partly predicted that in the liver. The present study demonstrates the complex organ-specific responses of postprandial fatty acid metabolism contributing to ectopic fat deposition and lipotoxicity during the development of cardiometabolic diseases.

## Data Availability

All data produced in the present study are available upon reasonable request to the authors.

## Acknowledgments

The authors would like to thank the participants of this study for their commitment and collaboration. The authors thank Caroll-Lynn Thibodeau, Maude Gérard, Myriam Flipot, Jennefer-Ann Lefrancois, Eric Lavallée, Esteban Espinosa, and Etienne Croteau for their excellent technical assistance and Stephen C. Cunnane for the use of PMOD. This study is part of the TIMED consortium which was supported by The Netherlands Organization for Health Research and Development (ZonMw) [459001021], Dutch Diabetes Research Foundation (Diabetes Fonds) [2019.11.101], Canadian Institutes of Health Research (CIHR) [TNC-174963], and Health-Holland [LSHM20107]. This collaborative project is co-financed with PPP-allowance made available by Health-Holland, Topsector Life Sciences & Health, to stimulate public-private partnerships.

## Authors contribution

Conceptualization, D.P.B. and A.C.C.; investigation, E.L.R., S.H.-L., M.L., L.D., F.F., G.R., C.N., W.M., S.P., M.F., L.B., B.G., É.E.T., A.C.C., and D.P.B.; visualization, E.L.R., S.H.-L., and D.P.B.; writing - original draft, E.L.R., S.H.-L., and D.P.B.; writing - review and editing, E.L.R., S.H.-L., L.D., C.N., O.S., W.M., G.R., B.G., É.E.T., F.R., P.R., JP.D., J.H., P.S., A.C.C., and D.P.B.; funding acquisition, A.C.C., D.P.B., F.R., P.R., JP.D., J.H., P.S..

---

**Supplemental Figure 1:**
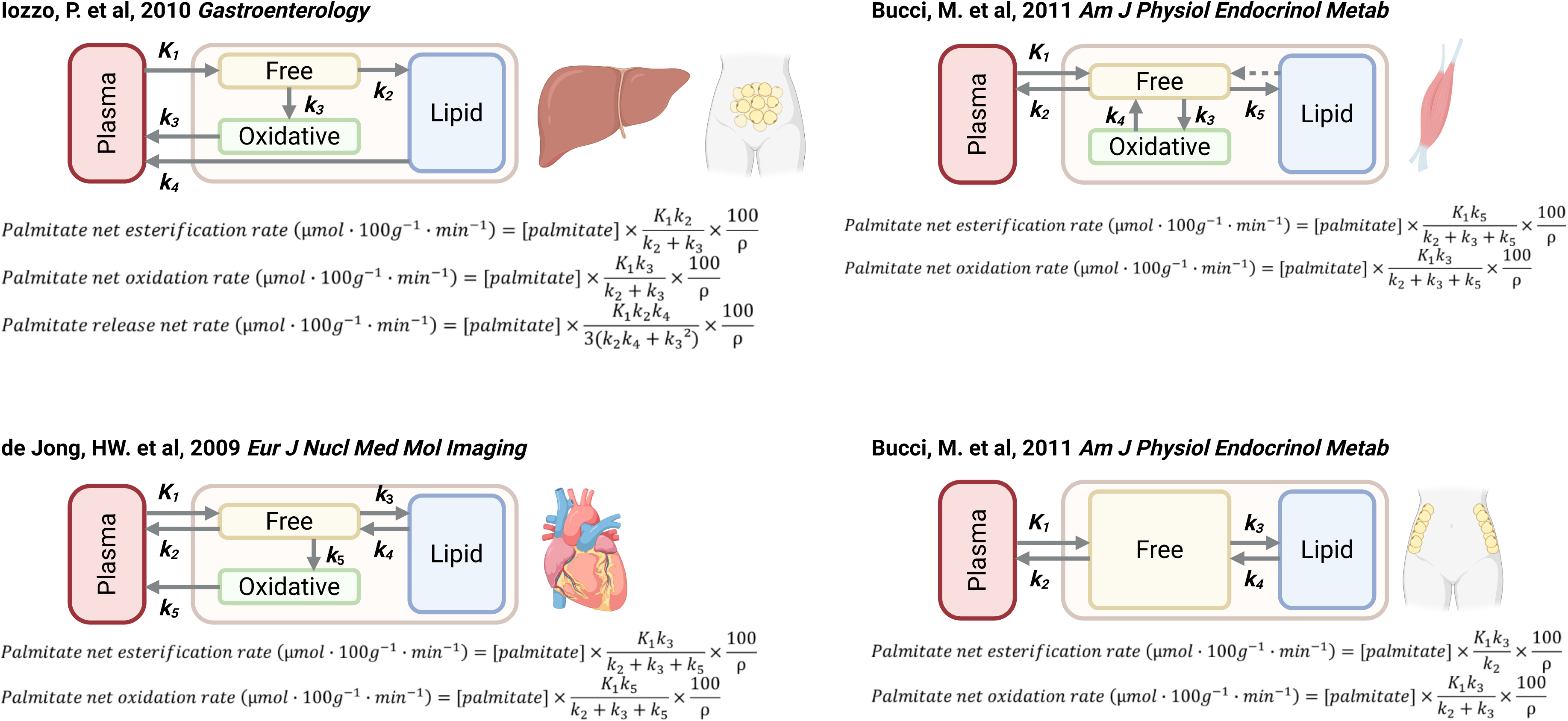
Presentation of the multi-compartmental models applied.

**Supplemental Figure 2:**
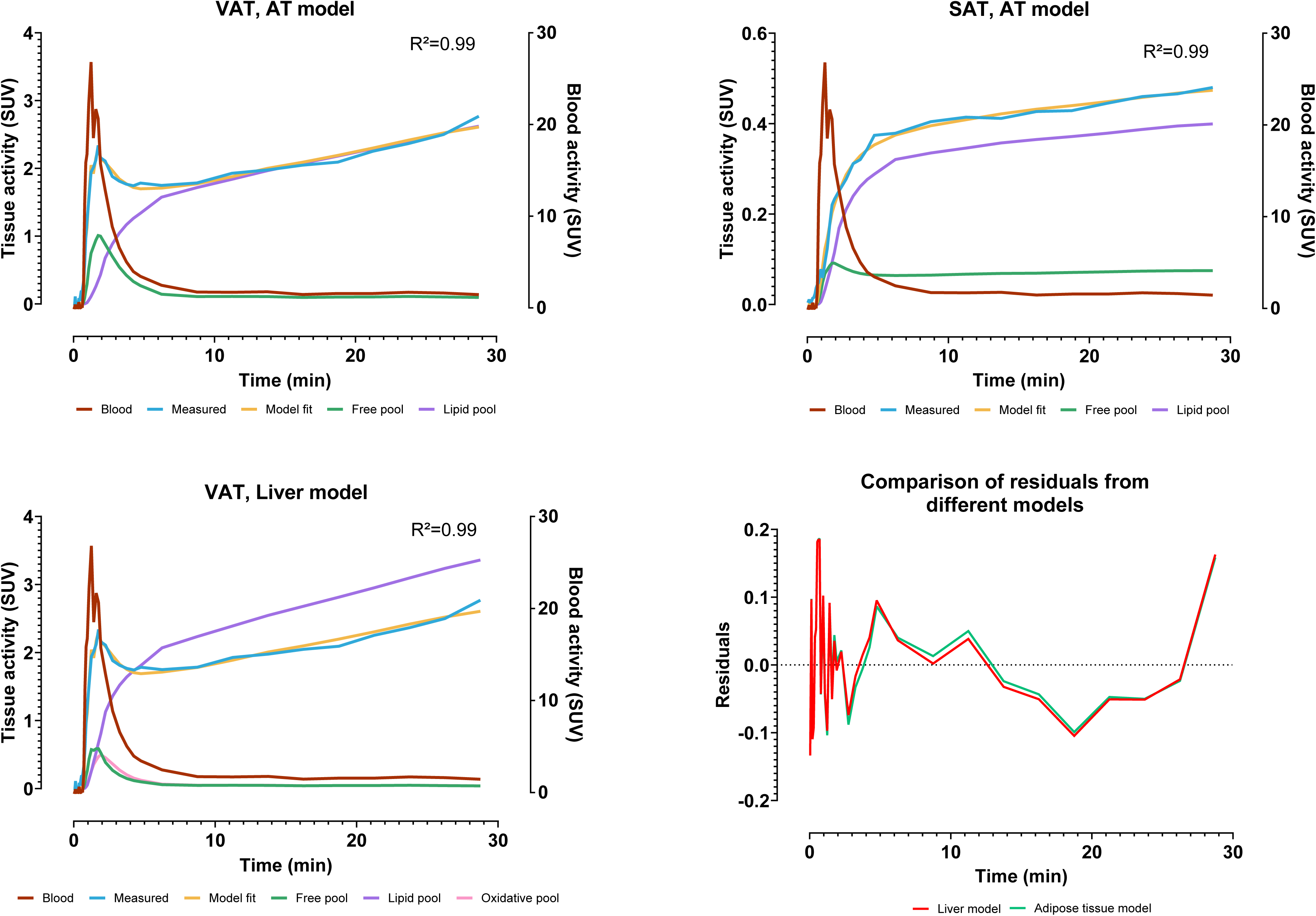
Comparison of adipose tissue and liver model for visceral adipose tissue modeling.

**Supplemental Table 1:** Tissue-specific kinetic parameters of palmitate metabolism in the fasting and postprandial states.

|  | Fasting | Postprandial | Linear mixed model analyses |  |
| --- | --- | --- | --- | --- |
|  | Lsmeans (95%CI) | Lsmeans (95%CI) | Estimated difference (95%CI) | P-Value |
| *Liver<sup>a</sup>* |  |  |  |  |
| $K_1$ (mL·mL<sup>-1</sup>·min<sup>-1</sup>)<sup>e</sup> | 0.34 (0.32 to 0.37) | 0.44 (0.41 to 0.48) | 30 (21 to 39) | <0.0001 |
| $k_2$ (min<sup>-1</sup>)<sup>e</sup> | 1.43 (1.17 to 1.75) | 2.27 (1.86 to 2.77) | 59 (23 to 105) | 0.001 |
| $k_3$ (min<sup>-1</sup>) | 1.05 (0.85 to 1.26) | 1.34 (1.14 to 1.54) | 0.29 (0.07 to 0.51) | 0.012 |
| $k_4$ (min<sup>-1</sup>) | 0.025 (0.021 to 0.029) | 0.038 (0.034 to 0.042) | 0.013 (0.010 to 0.017) | <0.0001 |
| *Skeletal Muscle<sup>b</sup>* |  |  |  |  |
| $K_1$ (mL·mL<sup>-1</sup>·min<sup>-1</sup>)<sup>e</sup> | 0.060 (0.050 to 0.072) | 0.051 (0.042 to 0.061) | -15 (-33 to 9) | 0.194 |
| $k_2$ (min<sup>-1</sup>)<sup>e</sup> | 1.13 (0.86 to 1.47) | 1.20 (1.03 to 1.40) | -7 (-16 to 37) | 0.570 |
| $k_3$ (min<sup>-1</sup>)<sup>e</sup> | 0.69 (0.44 to 1.09) | 1.23 (0.79 to 1.91) | 77 (-6 to 234) | 0.073 |
| $k_4$ (min<sup>-1</sup>)<sup>e</sup> | 0.41 (0.21 to 0.79) | 0.24 (0.17 to 0.34) | -43 (-71 to 13) | 0.103 |
| $k_5$ (min<sup>-1</sup>)<sup>e</sup> | 0.15 (0.10 to 0.22) | 0.25 (0.17 to 0.36) | 67 (2 to 174) | 0.042 |
| *Heart<sup>c</sup>* |  |  |  |  |
| $K_1$ (mL·mL<sup>-1</sup>·min<sup>-1</sup>) | 0.29 (0.27 to 0.30) | 0.37 (0.34 to 0.39) | 0.08 (0.06 to 0.11) | <0.0001 |
| $k_2$ (min<sup>-1</sup>) | 0.13 (0.09 to 0.17) | 0.06 (0.04 to 0.08) | -0.07 (-0.12 to -0.02) | 0.005 |
| $k_3$ (min<sup>-1</sup>)<sup>e</sup> | 0.22 (0.14 to 0.34) | 0.06 (0.04 to 0.09) | -73 (-85 to -51) | <0.0001 |
| $k_4$ (min<sup>-1</sup>)<sup>e</sup> | 0.46 (0.32 to 0.67) | 0.26 (0.12 to 0.54) | -44 (-75 to 28) | 0.165 |
| $k_5$ (min<sup>-1</sup>) | 0.18 (0.15 to 0.20) | 0.24 (0.19 to 0.28) | 0.06 (0.02 to 0.10) | 0.003 |
| *Visceral adipose tissue<sup>a</sup>* |  |  |  |  |
| $K_1$ (mL·mL<sup>-1</sup>·min<sup>-1</sup>)<sup>e</sup> | 0.12 (0.10 to 0.15) | 0.13 (0.11 to 0.16) | 7 (-16 to 36) | 0.565 |
| $k_2$ (min<sup>-1</sup>)<sup>e</sup> | 0.75 (0.59 to 0.94) | 0.95 (0.75 to 1.20) | 27 (-3 to 67) | 0.077 |
| $k_3$ (min<sup>-1</sup>) | 3.58 (2.65 to 4.51) | 4.18 (3.24 to 5.15) | 0.60 (-0.64 to 1.84) | 0.334 |
| $K_4$ (min<sup>-1</sup>) | 0.046 (0.039 to 0.052) | 0.018 (0.011 to 0.025) | -0.028 (-0.035 to -0.021) | <0.0001 |
| *Subcutaneous adipose tissue<sup>d</sup>* |  |  |  |  |
| $K_1$ (mL·mL<sup>-1</sup>·min<sup>-1</sup>) | 0.008 (0.006 to 0.010) | 0.010 (0.008 to 0.011) | 0.002 (-0.000 to 0.004) | 0.094 |
| $k_2$ (min<sup>-1</sup>)<sup>e</sup> | 0.64 (0.46 to 0.91) | 0.51 (0.36 to 0.72) | -21 (-51 to 27) | 0.315 |
| $k_3$ (min<sup>-1</sup>)<sup>e</sup> | 0.51 (0.35 to 0.74) | 0.65 (0.45 to 0.95) | 27 (-25 to 117) | 0.361 |
| $k_4$ (min<sup>-1</sup>) | 0.19 (0.03 to 0.35) | 0.08 (0.06 to 0.11) | -0.11 (-0.26 to 0.05) | 0.174 |

Data are estimated least-square means (LSmeans; 95% CI) and estimated within-subject differences (95% CI) from linear mixed-effects models accounting for repeated measures, with nutritional state as fixed effect and subject as random effect.
Multicompartmental models used: <sup>a</sup>Iozzo, P. et al, 2010, Gastroenterology; <sup>b</sup>Bucci, M. et al, 2011, Am J Physiol Endocrinol (skeletal muscle); <sup>c</sup>de Jong, L.J. et al, 2009, Eur J Nucl Med Mol Imaging; <sup>d</sup>Bucci, M. et al, 2011, Am J Physiol Endocrinol (adipose tissue).
<sup>e</sup>Statistics are performed on log-transformed variables; back-transformed LSmeans (95% CI) and model-derived fasting-to-postprandial effects are expressed as percentage change, derived from the exponentiated contrast of the linear mixed-model on the log scale, with 95% CI obtained by back-transforming the corresponding confidence limits are presented.

